# Genome-wide association and multi-omics functional screens reveal the genetic architecture of foveal development

**DOI:** 10.64898/2026.06.11.26355452

**Authors:** Callum Hunt, Manjiri Patil, Hammad Syed, Ha-Jun Yoon, Tingting Yang, Vanessa Rodwell, Zhanhan Tu, Gail DE Maconachie, Kayesha Coley, Alvin Lirio, Nick Shrine, Richard Packer, Mahmoud Fassad, Riddhi Shenoy, Natalie Allcock, Brandon Lim, Helen J Kuht, Girish Varma, Irem Karaer, Ranjit Injety, William Jakins, Reenette Savant, Rishi Sekhri, Michael Hisaund, Jinu Han, Seema Teli, Jun Wang, Zhen Zuo, Josh Whittingham, Gareth Douglas, Nicolas Sylvius, Pradeep C Vasudevan, Ala Moshiri, Jonathan H. Zippin, Brian P. Brooks, Lluis Montoliu, Irene Gottlob, Ko-Fan Chen, Takeshi Yoshimatsu, Martin D Tobin, William HJ Norton, Rui Chen, Chiara Batini, Mervyn G. Thomas

**Affiliations:** Ulverscroft Eye Unit, School of Psychology and Vision Sciences, University of Leicester, Leicester, UK; Department of Neurology, The Johns Hopkins University School of Medicine, Baltimore, MD 21287, USA; Department of Molecular and Human Genetics, Baylor College of Medicine, Houston, TX, USA; The School of AHPPNM, Faculty of Health, Division of Ophthalmology and Orthoptics, The University of Sheffield, Sheffield, United Kingdom; Division of Public Health and Epidemiology, University of Leicester, Leicester, UK; University Hospitals of Leicester NHS Trust, Leicester, UK; Electron Microscopy Facility Core Biotechnology Services, College of Life Sciences, University of Leicester, Leicester, UK; International Institute of Information Technology Hyderabad, Gachibowli, Telangana, India; Department of Ophthalmology, University Hospitals of Leicester NHS Trust, Leicester, United Kingdom; Institute of Vision Research, Department of Ophthalmology, Yonsei University College of Medicine, Seodaemun-gu, Seoul, Korea (the Republic of); School of Engineering, University of Leicester, Leicester, UK; NUCLEUS Genomics, Core Biotechnology Services, University of Leicester, Leicester, UK; Department of Ophthalmology & Vision Science, University of California Davis School of Medicine, Sacramento, California, United States; Department of Dermatology, Weill Cornell Medical College of Cornell University, New York, New York, United States; Ophthalmic Genetics and Visual Function Branch, National Eye Institute, National Institutes of Health, Bethesda, Maryland; Department of Molecular and Cellular Biology, National Centre for Biotechnology (CNB-CSIC), Madrid, Spain; Centre for Biomedical Network Research on Rare Diseases (CIBERER-ISCIII), Madrid, Spain; Neurology, Cooper University Health Care, Camden, New Jersey, United States; Division of Genetics and Genome Biology, University of Leicester, Leicester, UK; Department of Ophthalmology and Visual Sciences, Washington University in St Louis School of Medicine, St. Louis, USA; The University of Leicester, Division of Biosciences Education, University Rd, Leicester; Brunson Center for Translational Vision Research, Gavin Herbert Eye Institute, Department of Ophthalmology and Visual Sciences, University of California, Irvine, School of Medicine, Irvine, CA, USA; Human Genome Sequencing Center, Baylor College of Medicine, Houston, TX, USA; Department of Biochemistry and Molecular Biology, Baylor College of Medicine, Houston, TX, USA

**Author notes:** These authors contributed equally to this work (Callum Hunt, Manjiri Patil, Hammad Syed, Ha-Jun Yoon, Rui Chen, Chiara Batini, Mervyn G. Thomas).

## Abstract

Foveal hypoplasia causes visual impairment across congenital eye disorders, yet the genetic programmes governing foveal development remain poorly characterised and no tractable model exists for foveal disease. In the first genome-wide association study of foveal hypoplasia, we identified 42 sentinel variants mapping to 54 effector genes supported by ≥ 2 criteria from a variant-to-gene framework incorporating developmental multi-omics. Disruption of six effector genes using mutant lines and CRISPR knockouts in the zebrafish high acuity zone recapitulates structural, functional, and ultrastructural hallmarks of foveal hypoplasia, establishing the first vertebrate disease model. Integration with human foetal single-cell and spatial transcriptomics reveals two temporal waves of effector gene expression and identifies Müller glia as critical mediators of foveal patterning. Phenome-wide analyses reveal foveal variants are pleiotropic with refractive, lenticular, and metabolic traits, connecting foveal development to anterior segment and systemic disease biology. These findings should inform mechanistic studies of macular disease.

## Introduction

High-acuity vision in humans depends critically on the fovea, a specialised region of the central retina distinguished by dense cone photoreceptor packing and a characteristic pit-like depression. Foveal development is a precise, multi-stage process involving the centrifugal displacement of inner retinal neurons, elongation of foveal cone outer segments, and establishment of a foveal avascular zone.^1–4^ Although this anatomic sequence has been well described, the underlying molecular events, both during embryogenesis and throughout postnatal maturation, remain poorly characterised.

Disruption of the foveal developmental program results in foveal hypoplasia (FH), a condition marked by reduction or absence of these cone specialisations and foveal pit and significantly reduced visual acuity.^5, 6^ FH is a hallmark of several congenital retinal disorders, including albinism, and represents a significant cause of childhood vision impairment.^7, 8^ Understanding the genetic and molecular basis of foveal development is therefore a clinical priority, yet progress has been constrained by two fundamental challenges: a paucity of genes that can explain normal and abnormal foveal development, and a lack of experimentally tractable model systems with which to investigate disease mechanisms.

We have made steps toward addressing the first of these challenges by conducting the first genome-wide association study (GWAS) of foveal morphology,^9^ but the translation of prioritised genes to clinical FH remained unclear. To address the second challenge, recent work has identified a specialised region in the temporal retina of zebrafish (*Danio rerio*), the high acuity zone (HAZ), that recapitulates some architecture of the cone-rich primate fovea,^10^ offering an unprecedented opportunity to interrogate genetic drivers of foveal development in a high-throughput vertebrate system.^10, 11^

Motivated by these efforts, we conducted the first GWAS of FH and functionally validated selected target genes in a zebrafish model of human foveal development. Together with developmental multi-omics to assess transferability and pleiotropy analyses for broader disease implications, we aimed to unravel the genetic architecture defining normal foveal development and FH.

## Methods

### Foveal hypoplasia in the UK Biobank Cohort

FH status was classified in UK Biobank (UKB) participants using optical coherence tomography (OCT) 2D B-scan images which passed cross-sectionally through the fovea, obtained using TOPCON Advanced Boundary Segmentation.^12^ B-scans were flattened to standardise across images and then processed to develop a supervised deep learning model using Amazon Rekognition (https://aws.amazon.com/rekognition/). A total of 2500 input images were used to train a binary classification Amazon Rekognition model, 1800 of which were labelled as “FH” and 700 of which were labelled “no FH”. The grading of foveal hypoplasia (FH versus no FH) was performed by three clinical experts using the Leicester Foveal Hypoplasia Grading System,^6^ employing a consensus-based approach to select only high-quality B-scans. We used the trained deep learning model to classify FH status in the full complement of ∼80,000 OCT scans available for each eye. FH status was then determined for each participant by filtering all image classifications with ≥ 95% confidence and checking for bilateral concordance. This threshold was selected based on a sensitivity analysis, which demonstrated that the ≥ 95% cutoff yielded optimal diagnostic performance, achieving a maximum sensitivity of 100.0%, specificity of 96.2%, and Youden’s index of 0.96 (**Supplementary Fig. 1**).

### Genome-wide association study

We selected individuals of White British ancestry (Data-Field 22006) with FH classifications for association testing (4,388 cases and 29,990 controls), ensuring none were flagged for genomic analysis exclusion (Data-Field 22010). We performed genome-wide association testing using REGENIE V3.2.3.^13^ For step 1 we used genotype array data which was filtered to exclude variants with > 10% missingness and to include variants with minor allele frequency (MAF) 0.01 and Hardy-Weinberg equilibrium (HWE) *P*-value > 1×10^−15^. For association testing in step 2 we utilised Genomics England (GEL) imputed genetic data (UKB Data-Field 21008), employing an approximate Firth logistic regression approach under an additive genetic model. Age, weight, height, sex and the first 20 genetic principal components were included as covariates. We filtered association results for INFO > 0.3 for variants with MAF ≥ 0.01 or INFO > 0.8 for variants with MAF < 0.01. Both the genomic control lambda and the linkage disequilibrium (LD) score intercept were calculated using LD score regression (LDSC) v1.0.1.^14^

#### Identifying sentinel variants

Variants with genome-wide significant association (P < 5×10^−8^) were grouped in 2 Mb loci for fine-mapping centred on the most significant variant in the locus. Fine mapping was then performed using SuSiE integrated within Polyfun^15^ V1.0. LD matrices for Polyfun were produced for each locus of interest using LDstore2,^16^ excluding genetic variants with a missingness data rate greater than 10% and using a reference sample of 10,000 unrelated White British participants (Data-Field 22006) in UKB. In cases where SuSiE failed to produce a credible set, we performed conditional analysis using GCTA-COJO^17^ V1.94 (--cojo-slct) and produced 95% credible sets for each independently associated variant using the Wakefield method,^18^ setting the prior W to 0.04. Sentinel variants were obtained from all credible sets, defined as the variant within the credible set with the highest individual posterior inclusion probability (PIP).

#### Novelty of sentinel variants

We assessed the novelty of our sentinel variants against six published GWAS studies of retinal layer thickness.^19–24^ We determined that our sentinels were novel if they were not in LD with any previously published associations (r^2^ < 0.1), calculated using PLINK 1.9 ^25^ in the GEL imputed dataset. In cases where the previously reported sentinel variants were not present within the GEL imputed dataset (UKB Data-Field 21008), LDlink was used instead with a European reference population (https://ldlink.nih.gov/?tab=home).

### Identifying foveal hypoplasia effector genes

Sentinel variants were mapped to effector genes to investigate their function and gain insights into the biology of FH. We did so by utilising 9 variant-to-gene criteria and prioritising genes supported by at least two of these criteria.

#### Nearest gene

The closest protein-coding gene to each sentinel variant was determined based on the distance to the transcription start site (TSS) of canonical transcripts, using Ensembl (v112) annotations accessed via the biomaRt package.^26^

#### Genic annotation

Variants were annotated as falling within the exonic, promoter, or untranslated region (UTR) regions of genes using the *ChIPseeker* R package ^27^ with *TxDb.Hsapiens.UCSC.hg38.knownGene*. Promoters were defined as ±1,000 bp from the TSS. All annotations were confirmed by manual inspection in the UCSC Genome Browser (hg38).

#### Cis-regulator element annotation in human developmental multi-omics data

Utilising a dual-omics human foetal dataset,^28^ we identified Open Chromatin Regions (OCRs) and Differentially Accessible Regions (DARs) from retinal single-nucleus ATAC-seq data using the ArchR framework.^29^ DARs were called using getMarkerFeatures() and getMarkers() (false discovery rate [FDR] ≤ 0.01, logarithm base-2 fold change ≥ 0.5). OCR-gene links were defined by correlating chromatin accessibility within ±250 kb of TSS with gene expression (getPeak2GeneLinks(), FDR ≤ 0.01, cor ≥ 0.5) and with promoter accessibility (getCoAccessibility(), cor ≥ 0.5). The union of both sets defined potential cis-regulatory regions.

#### Retinal eQTLs

We performed a direct lookup of GWAS sentinel variants within a comprehensive dataset of published human retinal expression quantitative trait loci (eQTLs).^30^ We prioritised eQTLs that met a genome-wide significance threshold of P < 5×10^−8^.

#### Blood pQTLs

We performed direct lookup of sentinel variants with three sources of blood protein quantitative trait loci (pQTLs) data: UKB (Olink); ^31^ deCODE Genetics (Somascan) ^32^ and the EXCEED study (mass spectrometry).^33^ For both UKB Olink and the EXCEED study we set the significance threshold at P < 5×10^−8^, but we used a threshold of P < 1.8×10^−9^ for DeCODE Genetics in line with the original publication. In each case we prioritised cis-pQTLs only (i.e., gene boundary is within ± 1Mb of the sentinel position) for variant-to-gene prioritisation.

#### Nearby mouse knockout orthologs with vision related phenotype

Human orthologs of mouse knockout (KO) genes with vision-related phenotypes were identified within ± 500 kb of sentinel variants (i.e., based on gene start or end boundaries). Mouse phenotype data were obtained from the MGI (https://www.informatics.jax.org/) database (phenotype term MP:0005391) and supplemented with manual PubMed searches of nearby genes.

#### Nearby Mendelian disease genes

We identified a list of relevant Mendelian diseases using Orphanet (https://www.orpha.net/) by searching the title and clinical signs of all diseases for the following search terms “fovea, macula, retin, albinism, pigment, photoreceptor, cone, Hermansky-Pudlak, idiopathic infantile nystagmus” and then obtained a list of all causative genes for the identified disorders. We selected resulting Mendelian disease genes among this list, which are within ± 500 kb of sentinel variants.

#### Polygenic priority score (PoPs)

Polygenic priority scores (PoPs)^34^ were computed for ∼18,000 protein-coding genes using FH GWAS summary statistics, incorporating 57,543 functional features from gene expression, protein–protein interaction, and biological pathway datasets. The gene with the highest PoPs within ± 500 kb of each sentinel variant was prioritised. We used the 1000 genome project (European; Phase 3) as the LD reference panel.^35^

#### Rare variant association

Rare-variant association testing was performed using whole-exome sequencing data (4263 FH cases and 28,978 controls) with REGENIE V3.4.1.^13^ Association testing was performed using the same REGENIE parameters as the GWAS, except we performed association testing for rare variants with MAF < 1% and minor allele count >3 and excluded variants recommended for exclusion by UKB. Second, we performed gene-burden testing with a set of published masks,^36^ using the REGENIE GENE_P omnibus test. We then sought to identify genes ± 500 kb of sentinel variants implicated by our rare-variant association analysis. To this end we used a suggestive threshold of P < 5×10^−6^ for single-variant tests and P < 2.24×10^−4^ for gene-burden tests reflecting a Bonferroni correction for the number of protein coding genes (n=223) ± 500kb of sentinel variants.

### Functional characterisation of effector genes

We investigated the likely role of effector genes for foveal development using GeneCards (https://www.genecards.org) and published literature in PubMed (https://pubmed.ncbi.nlm.nih.gov). We then performed pathway enrichment analysis using ConsensusPathDB (http://cpdb.molgen.mpg.de/).

### Fish maintenance and zebrafish strains

*Danio rerio* were maintained under standard conditions at the Pre-clinical Research facility at the University of Leicester under Project License PP1567795 and Establishment License X1798C4D2. Embryos were raised in an incubator at 28.5°C. Wild-type AB strains were used for all CRISPR KO experiments. Two mutant lines for *tyr* (tk20) and *oca2* (sa9009) were imported from EZRC (https://www.ezrc.kit.edu/catalog.php).

### CRISPR Cas9 F0 knockouts in zebrafish

Three loci were targeted for *ahr1b, cyp26a1, jarid2a, jarid2b* and two for *lhx2b* to generate F0 KOs using a previously described protocol.^37^ The synthetic guide RNAs (Supplementary Table 1) and green fluorescent protein-tagged Cas9 protein were obtained from Integrated DNA Technologies (https://eu.idtdna.com/page). KO was confirmed by Sanger Sequencing using gene specific primers (Supplementary Table 2).

### Optokinetic reflex assay in zebrafish

Visual behaviour of paired wildtype and F0 KO/mutant embryos was assessed at 5 days post fertilisation (dpf) using an infrared light set-up previously described.^38^ To estimate visual acuity (VA), the slow phase velocity at different spatial frequencies and the spontaneous eye drift at the stationary phase were calculated for statistical comparison. For *lhx2b* KO embryos, inclusion in subsequent analyses was restricted to those exhibiting a reversed optokinetic response, consistent with the phenotype observed in *belladonna* mutants.^39^ Differences between the wild-type and mutant group VA were analysed using a Kruskal-Wallis test, followed by Steel’s many-to-one rank test for post-hoc comparisons. Statistical significance was set at *P* < 0.05

### Micro-CT Imaging and Analysis

Formalin fixed 5dpf embryos were washed in PBS and placed in 70% ethanol for an hour before overnight staining in 0.3% PMA in 70% ethanol. After ethanol dehydration and propylene oxide wash, a graded series of PO : Spurr’s low viscosity resin was used to transition samples to 100% resin. The resin-embedded embryos were polymerised at 60°C for 16 hours using an adapted ‘drip’ embedding method^40^ and mounted on a carbon rod for imaging.

Imaging was performed on a ZEISS Xradia CrystalCT (Supplementary Table 3) with voxel size 1µm for all runs. Eyes were aligned with Fiji software in dorsal-ventral and anterior-posterior axes, keeping the position of optic fissure consistent. Images were resliced from sagittal volume starting from the superior retina, covering the whole eye in 1° increments. A custom-trained deep learning model was built on ZEISS ARIVIS-Cloud (www.arivis.cloud) for Pixel-based segmentation of different layers of the retina.

To calculate the thickness of the photoreceptor layer (PRL) at the zebrafish HAZ, resliced segmented images were reconstructed into a polar plot representing the entire retina. Maximum PRL thickness was calculated within the 100° and 140° polar angle range, corresponding to the high acuity zone, after excluding the top 1% of thickness values to reduce the influence of imaging artifacts and segmentation outliers. Differences in maximum PRL thickness between wild-type and mutant groups were compared using a Kruskal-Wallis test, with Steel’s many-to-one rank test for post-hoc comparisons against the wild-type control. Statistical significance was defined as P < 0.05.

### Array tomography

Wild-type and *tyr* mutant embryos were embedded following the protocol adapted from Schmitt and Dowling., 1999.^41^ Briefly, following primary fixation, 5 dpf embryos were washed in 0.1M cacodylate buffer, and incubated in osmium solution 1. With water wash steps, samples were treated with 1% Thiocarbohydrazide (TCH), stained with 1% osmium tetroxide, and incubated overnight in 1% uranyl acetate at 4°C. After ethanol dehydration and propylene oxide wash, a graded series of PO : TAAB resin was used for infiltration, followed by a 100% TAAB incubation overnight (Supplementary Table 4). Finally, samples were embedded in plastic moulds with fresh TAAB and polymerized for 16 hours at 60°C.

Serial sections of 70nm thickness were obtained using Leica EM UC7 ultramicrotome. Sections were collected onto glow discharged indium tin oxide (ITO) coverslips utilising the paperclip modification technique,^42^ and dried for 30 min at 70°C. Images were obtained at 60nm resolution on a Zeiss Gemini 360 SEM with ATLAS5 software and using a 4-ring annular backscatter electron (BSE) detector with all quadrants selected to inverted (Supplementary Table 5).

3D volumetric data was aligned using DragonFly (v2022.2.0.1409). For vertical view, 100 serial images were segmented via transfer learning using a Detectron2 model (modified from https://github.com/bnsreenu). The model was trained and validated on wild-type and *tyr* sections, with training and validation split of 123 and 31 respectively (∼80% training and ∼20% validation). The final obtained masks manually refined in DragonFly. The model achieved a mean Average Precision (mAP) of 64.6% at IoU thresholds of 0.50:0.95, and 80.6% at IoU 0.50.

Quantitative analysis on the masks was performed using Python and the scikit-image library. For each cellular object the cell’s area, perimeter, and Feret max diameter was measured. Objects with an area less than 20 pixels and touching boundaries were disregarded. Differences in these cone specialisation measurements between wild-type and *tyr* mutant groups were compared using a Wilcoxon rank-sum test with Bonferroni correction for multiple comparisons. Statistical significance was defined as P < 0.05.

### Transcriptomics datasets and analysis

For single-cell transcriptomic analysis, we utilised a published single-nucleus RNA sequencing atlas of the developing human neuroretina spanning foetal weeks 10-23.^28^ This atlas encompasses the major neuroretinal cell classes, including retinal progenitor cells, retinal ganglion cells, amacrine cells, horizontal cells, bipolar cells, cone photoreceptors, rod photoreceptors and Müller glia, but not retinal pigment epithelium. Visualisation of gene expression across cell types and developmental timepoints was conducted using Seurat ^43^ (Version 5.2.1).

For spatial transcriptomic analysis, we generated Visium HD data from fresh-frozen human foetal retinal tissue at 10 weeks gestation. While structural foveal specification is well-documented to begin around 20 weeks gestation,^1–4^ this early timepoint was utilised to capture the foundational molecular patterning and transcriptional gradients that precede morphological changes. This research complies with the tenets of the Declaration of Helsinki and was reviewed by the UC Davis Institutional Review Board (IRB ID: 903054-1). The use of ocular tissues from discarded de-identified human foetal material was approved by the UC Davis Stem Cell Research Oversight Committee (SCRO protocol #1171, initial approval 12/16/2019). Tissues were obtained from donors who freely agreed to allow foetal tissue to be used for research purposes; informed consent was documented in the medical chart. Patients were not compensated. Human samples were collected from the UC Davis Eye Center within 6 hours post-mortem.^44^ Whole globes were dissected free from extraocular tissues. For eyes at or below 14 weeks gestation, globes were flash-frozen without fixation in isopentane and embedded in optimal cutting temperature compound. For eyes beyond 14 weeks gestation, globes were fixed in 4% PFA/5% sucrose for 48 hours or in Modified Davidson’s Fixative for approximately 24 hours before transfer into 100 mM glycine as a fixative quencher. Fixed tissue was immersed in a sucrose gradient (10%, 20%, and 30%) for cryoprotection before optimal cutting temperature embedding. All tissues were derived from discarded de-identified human foetal material under HIPAA Privacy Rules.

For Visium HD spatial gene expression, fresh-frozen tissues were embedded in optimal cutting temperature and cryosectioned at 10 μm thickness at −15°C (Leica CM 1950). Sections were mounted onto Visium CytAssist Tissue Slides (10x Genomics Visium HD) and stored at −80°C until use. Tissue sections were fixed in methanol at -−20°C, stained with haematoxylin and eosin, and imaged using a 20x objective on a Keyence BZX800 microscope. Samples were destained before probe hybridisation, ligation, slide preparation, probe release, extension, library construction, and sequencing, following the Visium HD Spatial Gene Expression Reagent Kits User Guide (CG000685). Sequencing was performed on an Illumina NovaSeq 6000 with paired-end reads (151 cycles Read 1, 8 cycles i7, 8 cycles i5, 150 cycles Read 2). Raw sequencing data were processed using Space Ranger (https://github.com/10XGenomics/spaceranger; 10x Genomics) with alignment to the GRCh38 human reference genome. Spatial bins were filtered for quality, normalised, and annotated to retinal cell types using reference-based label transfer from the single-nucleus RNA sequencing atlas.^28^ Regional segmentation of the retina relative to the foveal centre was performed to quantify gene expression gradients from macula to periphery.

### Genetic correlation analysis

Using summary statistics from published genome-wide association studies in European individuals (Supplementary Table 6), we calculated genetic correlations between each trait and FH using LD Score regression (LDSC v1.0.1^14^); with pre-computed LD scores for European populations. We specifically focused on ocular traits relevant to foveal development and morphology.

### Phenome-wide association study

DeepPheWAS ^45^ was used to perform association testing between sentinel variants and 1939 phenotypes defined in UKB, adjusting for age, sex, genotyping array and 10 principal components of genetic ancestry with default parameters. Phenome-wide associations at FDR <1% are reported.

## Results

In this study we mapped the genetic architecture of FH by combining genomic discovery with functional validation in a multi-tiered framework presented in **Figure 1**.

**Figure 1.**
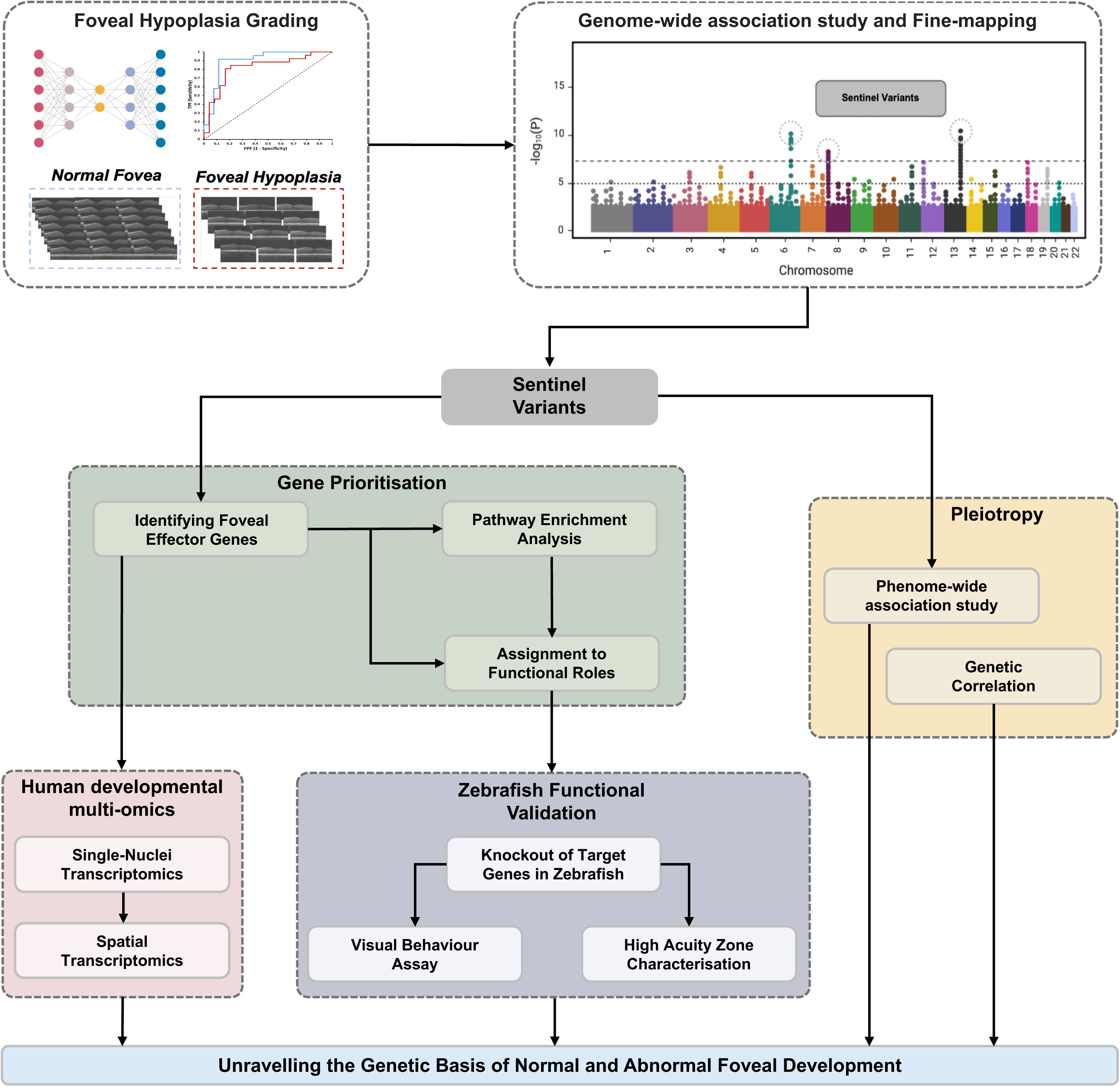
Study Design.

### Genome-wide association study of foveal hypoplasia identifies 54 effector genes

We utilised optical coherence tomography scans within UKB to classify FH using a supervised deep-learning approach based on clinical grading criteria.^6^ Our assembled cohort comprised 4,388 cases and 29,990 controls (**Fig. 2A**). Genome-wide association analysis of 29,459,844 variants (**Fig. 2B**) identified 42 sentinel variants after fine-mapping (Supplementary Table 7), of which 14 represent novel signals (Supplementary Table 8). The remaining 28 sentinels overlap with retinal layer thickness loci,^19–24^ consistent with FH involving altered laminar cell fate in addition to structural disruption. The median number of variants within 95% credible sets was nine. Eleven sentinels reached a posterior inclusion probability (PIP) ≥ 0.9, and 15 reached PIP ≥ 0.5.

**Figure 2:**
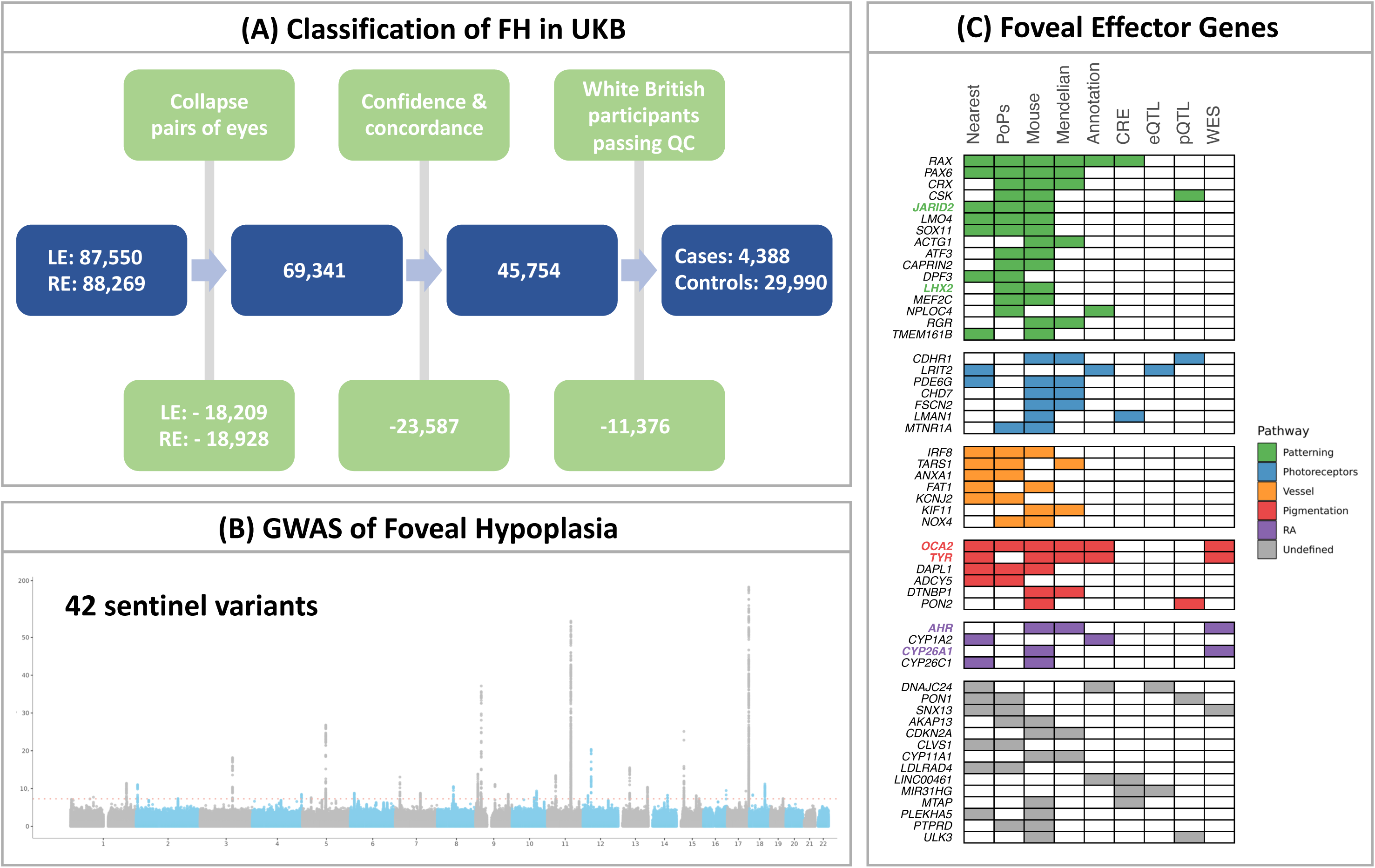
Gene prioritisation framework for FH. **(A)** Workflow for deep-learning phenotype definition. Scans were collapsed per participant, filtered for > 95% FH prediction confidence, and checked for bilateral agreement. QC involved restricting the sample to White British participants and excluding cases with recommended exclusion flags or sex discordance. **(B)** GWAS results identifying 42 sentinel variants post-fine-mapping; λ_QC_ =1.046; LDSC-intercept = 1.001. **(C)** Prioritisation of 54 effector genes. Plot indicates supporting evidence per gene, categorised by prioritised functional pathway. Emboldened and coloured genes were prioritised for downstream functional characterisation. FH: foveal hypoplasia; UKB: UK Biobank; GWAS: Genome-wide association study; PoPs: Polygenic priority score; CRE: cis-regulatory element; eQTL: expression quantitative trait loci: pQTL: protein quantitative trait loci; WES: whole-exome sequencing.

To map sentinel variants to effector genes, we deployed a multi-omics variant-to-gene framework incorporating variant annotation, cis-regulatory element mapping within human foetal developmental datasets, expression and protein quantitative trait loci, and rare-variant analysis. This approach prioritised 54 foveal effector genes with ≥ 2 lines of evidence (**Fig. 2C**, Supplementary Table 9-10). Of these, 23 were unique to FH when compared with our previously published foveal pit depth GWAS effector list.^9^ Strikingly, this set was dominated by cell fate and patterning determinants, including *ATF3*, *LMO4*, *SOX11*, *JARID2*, *LHX2*, *CRX*, *DPF3*, and *NPLOC4*. We also identified distinct novel genes involved in photoreceptor biology (*CHD7*, *MTNR1A*), pigmentation (*DTNBP1*, *PON2*), retinoic acid metabolism (*CYP26C1*, *CYP1A2*), and blood vessel development (*NOX4*, *KCNJ2*, *KIF11*, *ANXA1*). Notably genes related to cytoskeletal or extracellular matrix function were absent, consistent with our phenotypic focus on FH as a developmental patterning defect rather than a measure of gross foveal morphology.

For functional validation in zebrafish, we selected six foveal effector genes implicated by our GWAS to represent three key pathways in foveal development: pigmentation, retinoic acid metabolism, and temporal patterning (**Fig. 2C**). *TYR* and *OCA2* both encode proteins required for melanin biosynthesis and represent the most common genetic causes of oculocutaneous albinism, and consequently FH, in European and African populations, respectively;^46^ their inclusion served to benchmark our zebrafish model against known foveal disease biology. *AHR* encodes the aryl hydrocarbon receptor, recently identified as a Mendelian cause of infantile nystagmus syndrome with associated FH.^47^ Beyond established disease genes, our GWAS implicated *CYP26A1*, encoding a cytochrome P450 enzyme responsible for retinoic acid degradation, a pathway also supported by our previous GWAS of foveal pit depth^9^ and increasingly recognised as central to foveal specification.^48^ Finally, we selected two novel foveal genes *LHX2* and *JARID2*, encoding a LIM-homeodomain transcription factor critical for gliogenesis^49^ and a Polycomb repressive complex 2 component involved in temporal patterning of retinal progenitors,^50^ respectively. While *LHX2* function has been explored in zebrafish, where the orthologue *lhx2b* (also known as *belladonna*)^39^ has established roles in neural patterning, axon guidance and eye morphogenesis, its contribution to HAZ development has not previously been investigated.

### Disruption of foveal effector genes impairs zebrafish high acuity zone structure

To validate the role of these effector genes in foveal biology, we leveraged the zebrafish high acuity zone (HAZ), a region of localised retinal thickening in the inferotemporal retina driven by increased cone photoreceptor density and specialisation. Recent work has established that this region shares key features with the human fovea,^10, 11^ and we reasoned that loss of function in our prioritised candidates should disrupt HAZ specialisation if they contribute to human foveal development.

First, we performed Micro-CT imaging and polar plot analysis of wild-type retinas, confirming the regional HAZ specialisation, with thickening restricted to polar coordinates between 100° and 140° (median HAZ thickness = 25 μm; **Fig. 3A**). We then assessed the impact of each prioritised candidate on HAZ specialisation. For both *tyr* (tk20) and *oca2* (sa9009) we utilised established lines, but performed CRISPR-mediated KOs for *cyp26a1*, *lhx2b*, *ahr1b*, and *jarid2a/jarid2b* (double mutant), selecting paralogs with highest ocular expression based on published developmental atlases. Loss of function in nearly all candidates resulted in significant attenuation of HAZ specialisation (**Fig. 3B**). As expected, the pigmentation mutants *tyr* (median 15 μm, *P*=0.0008) and *oca2* (median 19 μm, *P*<0.0001) showed marked reductions, consistent with the established role of pigmentation in retinal development. Among the remaining candidates, the most severe phenotype was observed in *cyp26a1* (median 15.5 μm, *P*=0.0004), followed by *jarid2a/jarid2b* (median 16 μm, *P*=0.013) and *ahr1b* (median 21.5 μm, *P*=0.042). The *lhx2b* mutants showed a modest, non-significant reduction (median 23 μm, *P*=0.45). Topographic analysis confirmed that thinning was spatially restricted to the HAZ rather than affecting global retinal thickness (**Fig. 3C–E**).

**Figure 3.**
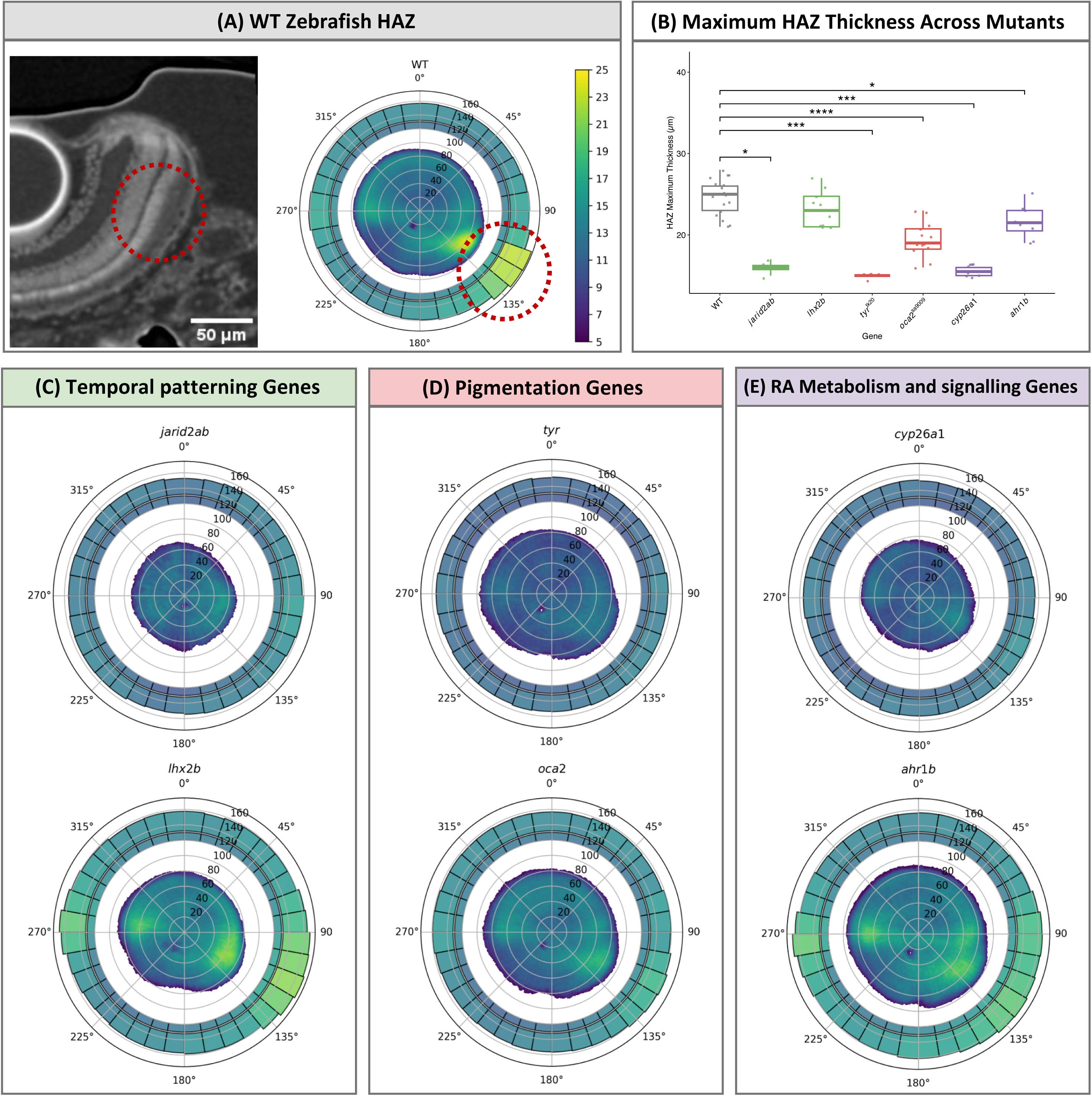
Micro-CT morphometry reveals localised retinal thinning in the zebrafish HAZ. **(A)** Representative micro-CT absorption contrast image of a WT zebrafish retina (left) with the HAZ indicated (orange circle). A corresponding polar plot (right) illustrates retinal thickness across the topographic map, with the HAZ region demarcated (orange circle). **(B)** Box plots showing maximum retinal thickness within the HAZ (coordinates 100°-140°) across indicated genotypes: WT(n=17 eyes), *jarid2a/jarid2b* (n=4), *lhx2b*(n=10), *tyr*(n=7), *oca2*(n=14), *cyp26a1*(n=8), and *ahr1b* (n=8). Statistical comparisons between WT and mutants were performed using Steel’s many-to-one rank test. Significance levels are denoted as ∗ *P*<0.05, ∗∗ *P*<0.01, *** *P* < 0.001, **** *P* < 0.0001. **(C-E)** Topographic polar plots demonstrate that retinal thinning in mutants is spatially restricted, peaking specifically within the HAZ (coordinates 100°-140°) when compared to WT controls. WT: Wildtype; HAZ; High-acuity zone.

### Structural deficits in the high acuity zone translate to impaired visual function

To determine whether the observed structural deficits in the mutant HAZ translate to functional visual impairment, we performed optokinetic response (OKR) assays, measuring the spatial frequency threshold at which larvae could no longer track moving stimuli. Wild-type larvae exhibited robust eye-tracking up to 0.134 cycles per degree (cpd), whereas tracking failure was observed in *jarid2a/jarid2b*, *oca2*, *tyr*, and *ahr1b* mutants at 0.08 cpd, and in *lhx2b* mutants at 0.10 cpd. (**Fig. 4A**). Plotting OKR slow-phase velocity against spatial frequency revealed constricted functional response ranges and premature reflex attenuation across mutant genotypes (**Fig. 4B-D**). Note that *cyp26a1* mutants were absent from this analysis as no eye-tracking response could be elicited at any frequency tested.

**Figure 4.**
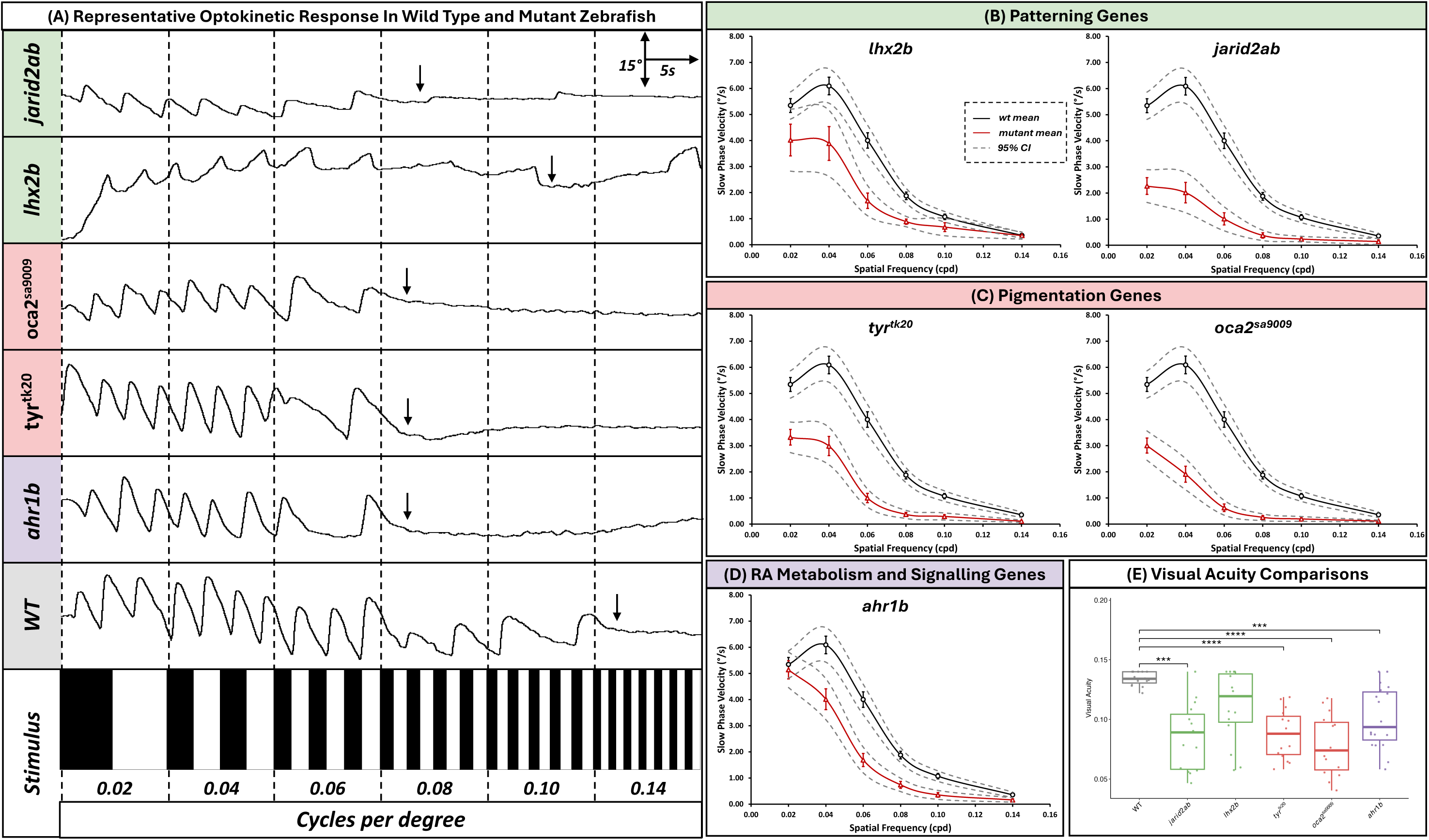
Mutant zebrafish exhibit impaired optokinetic response (OKR) and reduced visual acuity. **(A)** Representative OKR eye-tracking traces at threshold spatial frequencies. Tracking failure is observed in *jarid2a/jarid2b*, *oca2*, *tyr*, and *ahr1b* mutants at 0.08 cycles per degree (cpd), and in *lhx2b* mutants at 0.10 cpd, compared to WT (0.134 cpd). **(B–D)** OKR slow-phase velocity (SPV) as a function of spatial frequency. Mutant lines exhibit a constricted functional response range and premature reflex attenuation compared to WT. Data represent mean and 95% confidence intervals for WT and mutants **(E)** Visual acuity estimates are significantly reduced in mutants compared to WT. Statistical comparisons between WT and mutants were performed using Steel’s many-to-one rank test. Significance levels: ∗∗∗*P*<0.001, ∗∗∗∗*P*<0.0001. For all panels, WT (n = 15) , *ahr1b* (n = 19), *jarid2a/jarid2b* (n = 15), *lhx2b* (n = 15), *oca2* (n = 15), and *tyr* (n = 15). WT: wild-type; cpd: cycles per degree, CI: confidence interval.

Statistical comparison of visual acuity thresholds (**Fig. 4E**) mirrored earlier structural findings, with the most severe impairments seen in pigmentation mutants. Specifically, *oca2* mutants displayed the lowest acuity (median 0.074 cpd, *P*<0.0001) followed by *tyr* (median 0.088 cpd, *P*<0.0001). The temporal patterning mutant *jarid2a/jarid2b* also demonstrated significant functional deficits (median 0.089 cpd, *P*=0.0002), as did the metabolism gene *ahr1b* (median 0.094 cpd, *P*=0.0005). Consistent with structural findings, *lhx2b* mutants exhibited only a marginal, non-significant decrease in visual acuity (median 0.119 cpd, *P*=0.13).

### Loss of tyrosinase disrupts cone photoreceptor elongation and mosaic packing

To elucidate the cellular mechanisms underlying retinal thinning and visual acuity loss in mutant zebrafish, we employed high-resolution array tomography to evaluate the ultrastructural organisation of cone photoreceptors within the HAZ. This was motivated by the lack of specialised cone elongation and disrupted photoreceptor packing seen in Mendelian FH.^6, 8^ To this end, we used the *tyr* mutant as our representative model given its severe phenotype and the clinical prevalence of albinism-associated FH.

Analysis of vertical sections (**Fig. 5A-C**) revealed that *tyr* mutants exhibited significantly reduced cone dimensions, indicating a failure of photoreceptors to achieve the elongated morphology characteristic of foveal cone photoreceptors. This was evidenced by decreased median cross-sectional area (wild-type 10.26 μm^2^ vs. *tyr* 7.57 μm^2^, P=6.6×10^−149^), Feret diameter (wild-type 5.79 μm vs. *tyr* 4.46 μm, P=8.3×10^−301^), and perimeter (wild-type 14.16 μm vs. *tyr* 11.49 μm, P=1.1×10^−258^). Conversely, analysis of horizontal sections (**Fig. 5D-F**) revealed expanded cone profiles in mutants, indicative of disrupted mosaic packing, with increased median area (wild-type 1.05 μm^2^ vs. *tyr* 3.36 μm^2^, P=3.5×10^−6^), diameter (wild-type 1.75 μm vs. *tyr* 3.32 μm, P=6.2×10^−8^), and perimeter (wild-type 4.24 μm vs. *tyr* 8.01 μm, P=1.3×10^−7^). Together, these data demonstrate that loss of tyrosinase results in reduced photoreceptor elongation and disrupted cone mosaic packing within the HAZ, consistent with the phenotype observed in human FH.^1, 6^

**Figure 5.**
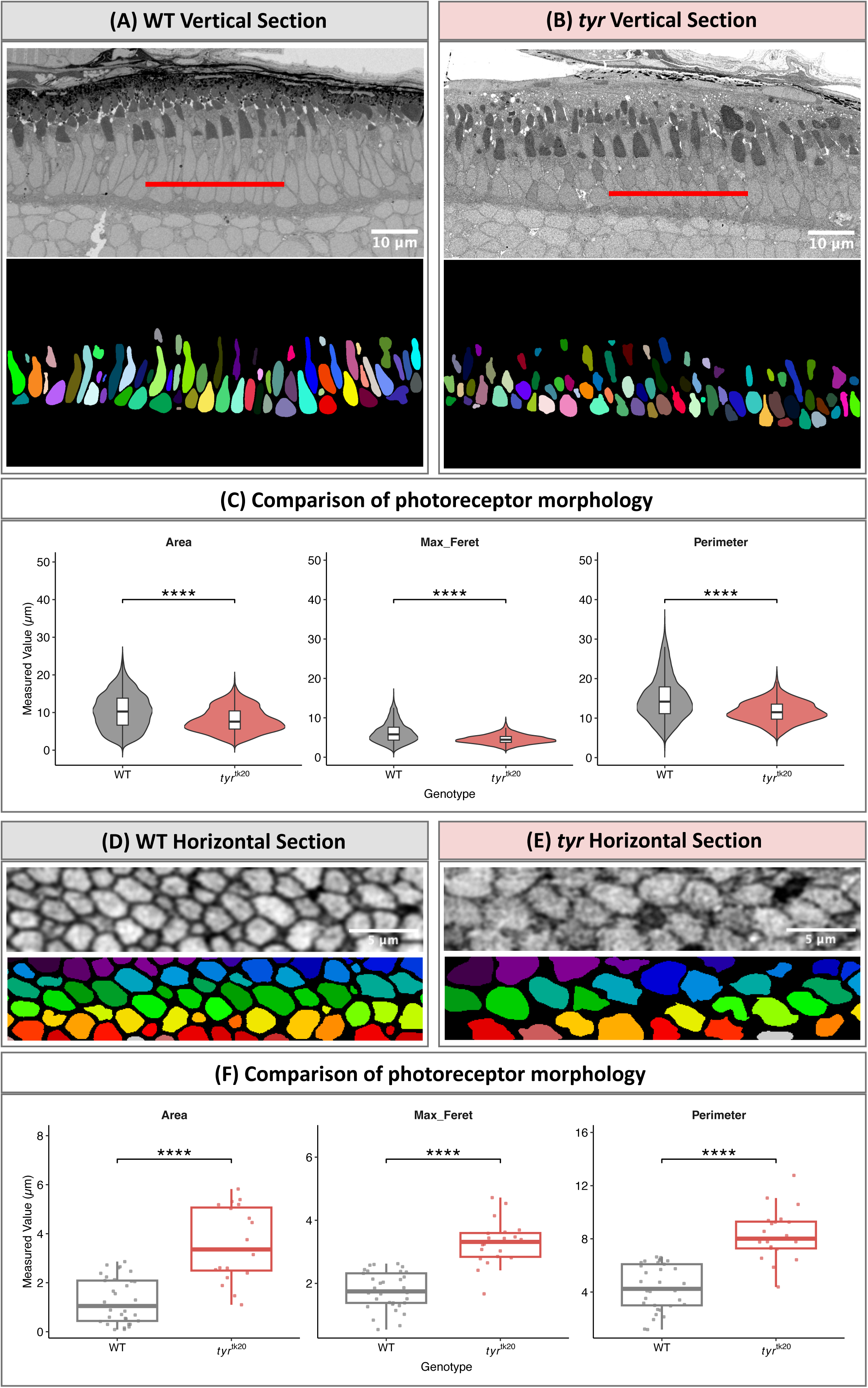
Volume electron microscopy (vEM) reveals impaired photoreceptor specialisation and packing in the *tyr* retina. **(A-B)** Representative vEM vertical sections of wild-type and mutant *tyr* retinas, with corresponding cone-mosaic projections. **(C)** Quantification of photoreceptor morphometry in vertical sections. *tyr* mutants exhibit significant reductions in cross-sectional area, Feret diameter, and perimeter, consistent with a failure of photoreceptor elongation and specialisation. **(D-E)** Representative horizontal sections and derived cone-mosaic maps of wild-type and mutant retinas. **(F)** Quantification of cell body metrics in horizontal sections. *tyr* mutants show reduced cell body area, diameter, and perimeter, indicating decreased cell density and disrupted mosaic packing. Statistical comparisons were performed using Wilcoxon rank-sum test. Significance levels: **** *P*<0.0001.

### Developmental multi-omics identify two waves of effector gene expression and implicate glia in foveal specification

To establish the developmental context of our findings and assess their translational relevance to human foveal biology, we intersected our 54 prioritised effector genes with a human multi-omics atlas spanning foetal weeks 10-23,^28^ the critical window during which foveal specification initiates histologically.^1^ Of the 54 effector genes, 53 were detected in the single-nucleus RNA sequencing dataset.

Temporal expression analysis across foveal and peripheral retina identified two distinct waves of effector gene activity (**Fig. 6A**). An early specification wave, prominent at 10-13 foetal weeks, was characterised by master regulators of retinal identity including *PAX6* and *RAX*, alongside *LHX2*, the novel effector gene identified by our study. A later specialisation wave, spanning 16-23 foetal weeks, was characterised by genes with higher expression in macular cell populations. This included established photoreceptor specialisation genes such as *CDHR1*, as well as the novel effector gene *CYP26A1 (***Fig. 6A**).

**Figure 6:**
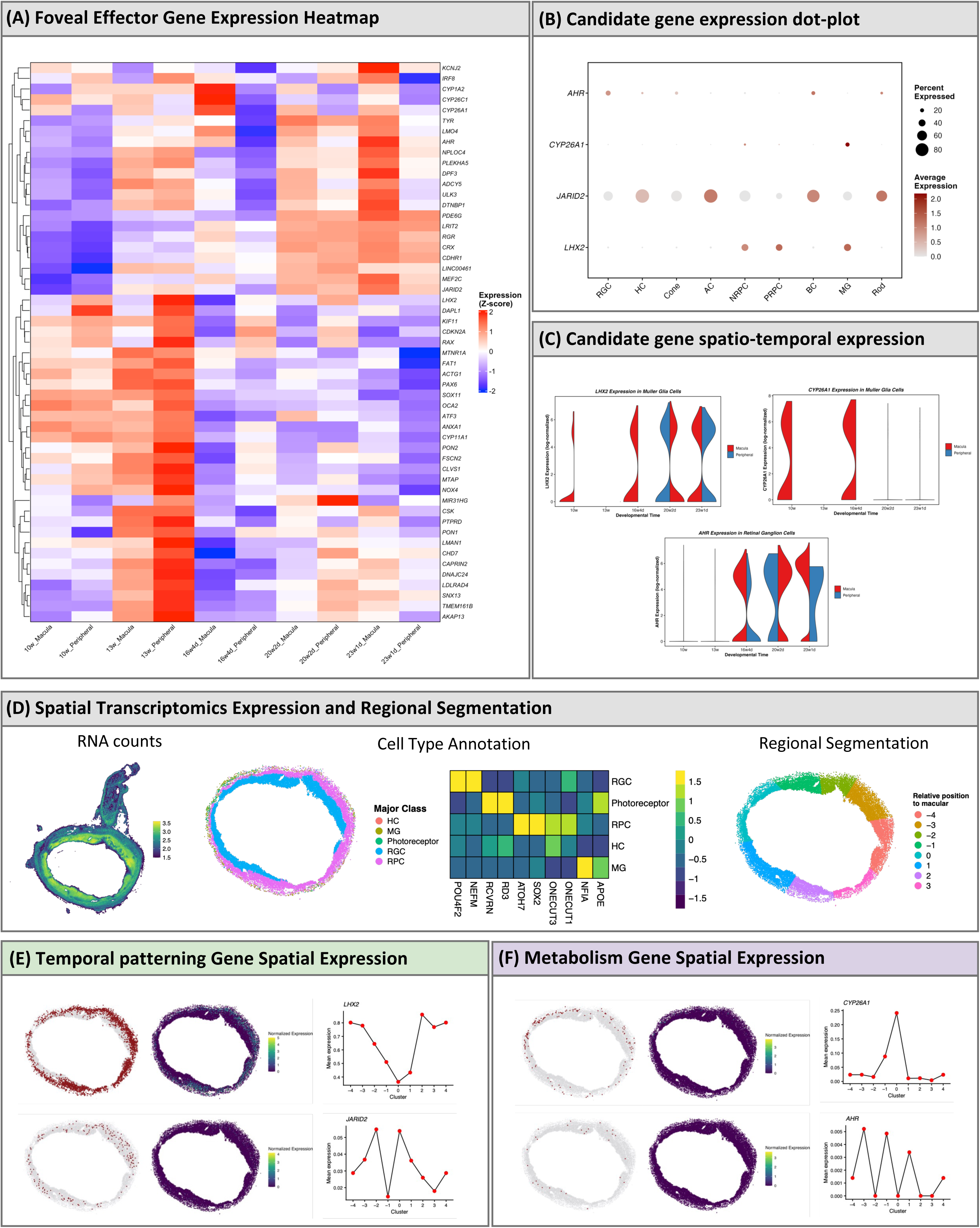
Single-cell and spatial transcriptomics analysis of foveal effector genes. **(A)** Heatmaps showing Z-score normalised aggregate gene expression in macula versus periphery from 10 to 23 foetal weeks. **(B)** Dot plot of aggregated cell-type expression for functionally validated candidate genes. Dot size indicates percentage of cells expressing the gene; colour intensity indicates average expression level. **(C)** Spatiotemporal expression of functionally validated target genes in key cell types: *LHX2* and *CYP26A1* in Müller glia, and *AHR* in retinal ganglion cells. **(D)** Workflow for the spatial transcriptomics pipeline, including RNA quantification, cell-type annotation and regional segmentation. **(E-F)** Spatial transcriptomics at 10 foetal weeks showing spatial expression of functionally validated genes. Red indicates expressing cells; adjacent panels show corresponding normalised expression. Line graphs display average expression at each segmented region relative to the macula (position 0) in retinal progenitor cells.

We next examined the cell-type expression profiles of functionally validated genes. Dot plot analysis revealed that *JARID2* was expressed broadly across multiple retinal cell types, varying in both the proportion of expressing cells and expression level (**Fig. 6B**). By contrast, *CYP26A1* and *LHX2* showed predominant expression in Müller glia, while *AHR* was detected in small populations of retinal ganglion and bipolar cells. Focused temporal analysis of these key cell types confirmed robust Müller glia expression for both *CYP26A1* and *LHX2*, with *CYP26A1* showing striking macular-specific enrichment during the 10-16 week developmental window (**Fig. 6C**). We observed that *AHR* expression in retinal ganglion cells was not regionally restricted and started at later developmental timepoints (16-23 weeks).

To resolve the spatial organisation of effector gene expression at higher resolution, we performed spatial transcriptomics on 10-week foetal retina (**Fig. 6D**). Spatial mapping of expressing cells revealed that *LHX2* and *JARID2* were detected broadly across the entire retina, whereas *CYP26A1* and *AHR* were expressed in isolated cell populations (**Fig. 6E-F**). Focused analysis of retinal progenitor cells across segmented retinal regions revealed distinct spatial gradients relative to the foveal centre, with *CYP26A1* demonstrating maximal expression at the fovea and *LHX2* displaying highest expression peripherally, whereas *AHR* and *JARID2* showed no regionally specific expression profiles (**Fig. 6E-F**).

### Foveal hypoplasia sentinel variants show extensive pleiotropy across ocular and systemic phenotypes

To evaluate the broader clinical implications of genetic variants associated with FH we conducted phenome-wide association analysis across 1,939 phenotypes in UKB (**Fig. 7A-B**, Supplementary Table 11).

**Figure 7:**
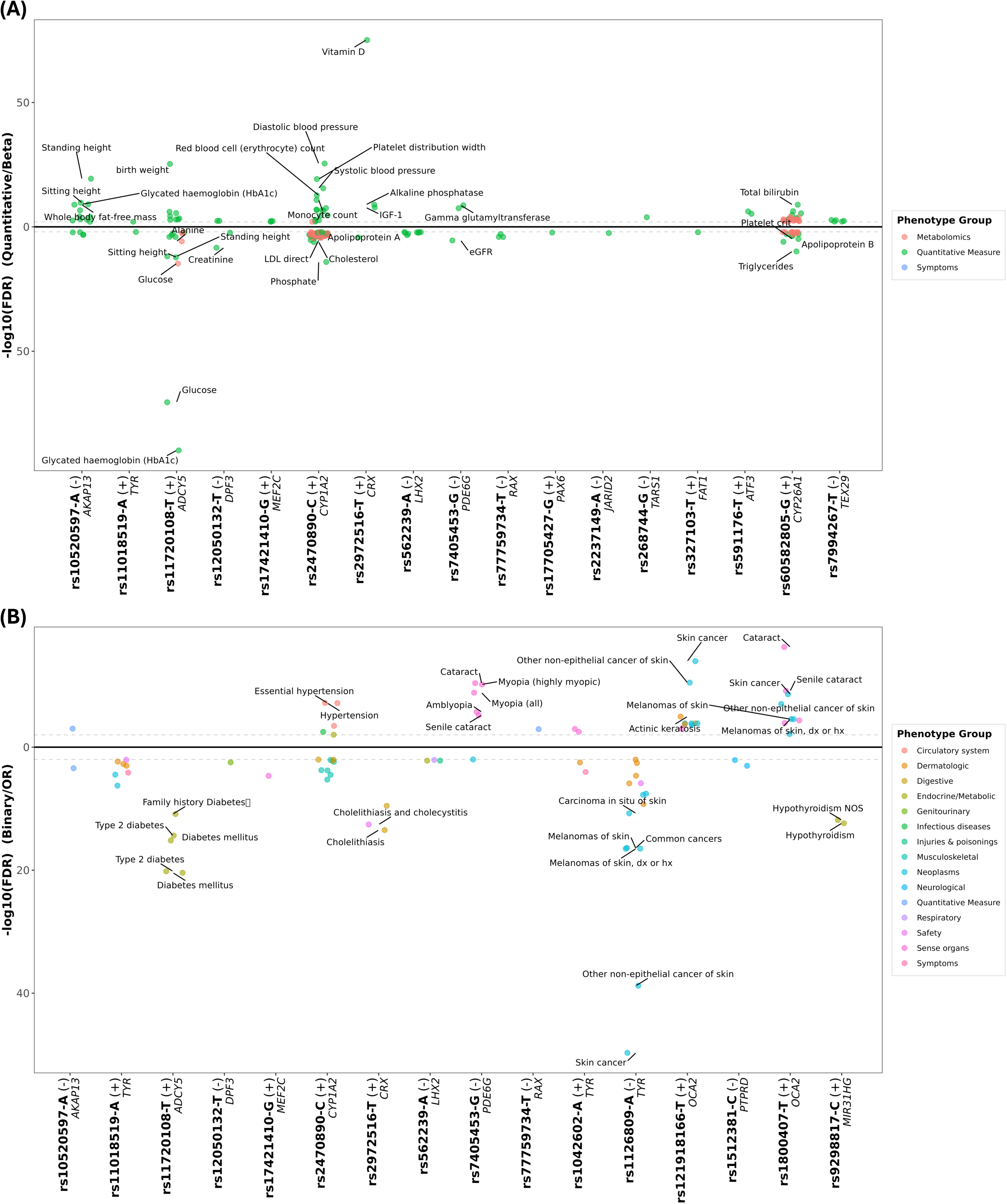
Phenome-wide association study of FH sentinel variants. (**A**) Associations with quantitative traits. (**B**) Associations with binary traits and case-control diseases. The x axis shows sentinel variants labelled by the GWAS effect allele and implicated gene, with the direction of effect from the foveal hypoplasia GWAS indicated (+ denotes OR > 1). The y axis shows -log_10_(FDR-adjusted *P*-value). Phenotypes are plotted above the zero line if their direction of effect is concordant with the GWAS effect, and below if discordant.

Foveal variants showed extensive pleiotropy with ocular conditions, particularly those affecting the lens and refractive development. The pigmentation variants *TYR* rs1042602-A and *OCA2* rs1800407-T were both associated with cataract (odds ratio [OR] range 1.03-1.12; FDR=3.3×10^−3^ and 5.3×10^−17^, respectively), with *OCA2* additionally associated with senile cataract (OR=1.13, FDR=6.9×10^−10^) and lens disorders (OR=1.17, FDR=4.5×10^−5^), and *TYR* with glaucoma (OR=1.07, FDR=1.1×10^−3^). The *PDE6G* variant rs7405453-G was also associated with cataract (OR=0.94, FDR=3.9×10^−11^), high myopia (OR=0.83, FDR=6.4×10^−11^), and amblyopia (OR=0.80, FDR=1.9×10^−6^), while a variant near *MEF2C* (rs17421410-G) was associated with myopia (OR=0.89, FDR=2.1×10^−5^). Despite this variant-level pleiotropy, genome-wide genetic correlation analysis revealed only modest and non-significant correlations between FH and other ocular conditions, including refractive error (r_g_ = −0.05), glaucoma (r_g_ = −0.02), age-related macular degeneration (r_g_ = −0.02), and diabetic retinopathy (r_g_ = −0.02), indicating that FH possesses a largely distinct genetic architecture. A strong negative correlation was observed with foveal pit depth (r_g_ = - 0.77, P = 6.8×10^−67^), consistent with FH representing the pathological extreme of reduced pit formation.

Foveal variants also showed associations with non-ocular phenotypes. Variants mapping to pigmentation genes showed associations with dermatological phenotypes, most notably *TYR* rs1126809-A with skin cancer (OR=1.14, FDR=2.0×10^−50^) and melanoma (OR=1.18, FDR=3.1×10^−17^). Several foveal variants also showed associations across metabolic traits: the *ADCY5* variant rs11720108-T was associated with glycated haemoglobin (β=-0.052, FDR=1.4×10^−90^), glucose (β=-0.050, FDR=3.0×10^−71^), and type 2 diabetes (OR=0.91, FDR=6.5×10^−21^), while the *CRX* variant rs2972516-T showed the strongest systemic association with serum vitamin D (β=0.058, FDR=8.0×10^−76^), alongside cholelithiasis (OR=0.89, FDR=3.3×10^−14^), IGF-1, and alkaline phosphatase. A *CYP1A2* variant (rs2470890-C) was associated with diastolic blood pressure (β=0.027, FDR=3.6×10^−26^), systolic blood pressure (β=0.020, FDR=3.2×10^−16^), hypertension (OR=1.04, FDR=6.8×10^−8^), and multiple lipid fractions including low-density and very low-density lipoprotein cholesterol and triglycerides. Anthropometric associations were limited to *AKAP13* rs10520597-A with standing height (β=-0.017, FDR=4.4×10^−20^) and *ADCY5* rs11720108-T with standing height (β=-0.014, FDR=1.5×10^−12^) and birth weight (β=0.037, FDR=5.1×10^−26^).

## Discussion

We conducted the first genome-wide association study of FH, identifying 42 sentinel variants mapping to 54 effector genes enriched for retinal patterning, photoreceptor specialisation, pigmentation, and retinoic acid metabolism. Functional validation in zebrafish demonstrated that disruption of prioritised candidates impairs HAZ structure and visual function, recapitulating human FH. Developmental multi-omics identified Müller glia as a key cell type in foveal development, and phenome-wide analyses revealed extensive pleiotropy linking foveal variants to broader ocular and systemic health.

The zebrafish HAZ recapitulates key features of the primate fovea, including a cone-dense region with specialised photoreceptor morphology and enhanced visual processing.^10, 51^ Previous work established that zebrafish exhibit striking temporo-nasal anisotropy in photoreceptor density and spectral tuning,^52^ with the inferotemporal strike zone displaying fovea-like cone specialisations that drive prey-capture behaviour.^10^ Building on this, we developed an integrated pipeline combining micro-CT morphometry, optokinetic response assays, and volume electron microscopy that faithfully models both normal foveal development and foveal disease. This pipeline provides a powerful framework for future functional interrogation of candidate genes at a scale not achievable in primate models, and for studies aimed at dissecting macular disease mechanisms and testing therapeutic interventions.

Utilising this system, we provide direct functional evidence that disruption of GWAS-prioritised genes impairs HAZ development, with severe structural and functional deficits observed across the mutant spectrum representing pigmentation (*tyr*, *oca2*), retinoic acid metabolism (*cyp26a1*), and temporal patterning (*jarid2*) pathways. CYP26A1-mediated retinoic acid degradation creates a localised zone of low retinoic acid signalling required for patterning high-acuity areas,^48^ and organoid studies have confirmed that this gradient shapes the cone subtype mosaic of the retina.^53^ JARID2, acting through Polycomb repressive complex 2, regulates the temporal progression of retinal progenitor competence, with loss of function extending early-born cell type production at the expense of later-born populations.^50, 54^ Together, these findings reveal an underappreciated role for temporal patterning and cell fate specification in foveal development, suggesting that disruption of signalling gradients or epigenetic temporal control impairs the coordinated cell type generation required for foveal patterning, potentially interacting with canonical pigmentation pathways to produce FH. In support of this, 28 of the 42 FH sentinel variants have been previously implicated in retinal layer thickness,^19–24^ further reinforcing the contribution of cell fate and patterning genes to foveal development.

Our developmental multi-omics analyses complement these functional data by revealing a two-wave temporal architecture of foveal effector gene expression. An early specification wave (10-13 foetal weeks) is characterised by master regulators of retinal identity including *PAX6*, *RAX*, and *LHX2*, whose broad expression across retinal progenitors establishes the regional framework upon which foveal identity is defined. A later specialisation wave (16-23 weeks) is dominated by genes acting in committed cell types, including the photoreceptor transcription factor *CRX* and the cone-specific cadherin *CDHR1*, alongside *CYP26A1* and *AHR*. This temporal separation mirrors the established sequence of primate foveal development, in which specification of foveal identity precedes the protracted process of cone packing, inner retinal displacement, and photoreceptor elongation that continues postnatally.^1, 3, 6, 55^ Notably, recent organoid work has demonstrated that *CYP26A1* itself exhibits biphasic expression in the presumptive macula, with an early wave promoting cell cycle exit and cone genesis and a later wave in Müller glia directing cone subtype specification towards a fovea-like composition.^56^ These findings have important therapeutic implications, since foveal maturation extends well beyond birth and the genes driving the specialisation wave may represent tractable targets for postnatal intervention.

In further support of a central role for Müller glia, cell-type expression analysis identified both *CYP26A1* and *LHX2* as showing expression in macular Müller glia populations. The macular-specific enrichment of *CYP26A1* during early development (10-16 weeks) aligns with evidence that retinoic acid suppression directs cone specification,^48^ supporting a model in which Müller glia contribute to foveal patterning through spatially restricted modulation of retinoic acid gradients. *LHX2* reinforces this glial involvement, since it is an essential factor for retinal gliogenesis, directly regulating Notch signalling and gliogenic transcription factor networks.^49^ However, *lhx2* KO had limited effect on zebrafish HAZ development, and spatial transcriptomics at 10 weeks of human foetal development confirmed that *LHX2* expression is highest in the peripheral retina, suggesting its gliogenic role may be more critical outside the foveal region. Nevertheless, Müller glia of the primate foveola have traditionally been considered structural elements,^57, 58^ and the convergence of two functionally validated effector genes on this cell type suggests a more direct role in foveal specification than previously recognised.

Phenome-wide analyses revealed broad implications of foveal variants for ocular and systemic health, with extensive pleiotropy observed for cataracts, myopia, and amblyopia. However, genome-wide genetic correlation analysis showed only modest correlations between FH and other ocular diseases, indicating that the observed pleiotropy is driven by specific shared loci rather than pervasive genetic overlap. The pleiotropy with cataracts and myopia suggests a developmental relatedness between the anterior segment and the posterior segment, specifically the fovea, that warrants further investigation. Associations with metabolic traits, including type 2 diabetes, glucose homeostasis, vitamin D, and lipid metabolism, may reflect the exceptionally high metabolic demands of the retina and its photoreceptors,^59, 60^ or shared developmental signalling pathways. The association with glucose and type 2 diabetes is particularly notable given that diabetes is an established risk factor for epiretinal membrane formation,^61^ a condition that directly affects macular structure. Anthropometric associations were limited to two loci, arguing against confounding by developmental maturity. Prematurity represents a major non-genetic cause of FH,^62–64^ and the absence of widespread anthropometric pleiotropy suggests that our GWAS captures intrinsic foveal developmental programmes rather than generalised growth effects.

We acknowledge limitations of our study. First our cohort was of European ancestry, limiting cross-ancestry inference, though functional follow-up supports the generalisability of discovered mechanisms. Future studies across diverse populations are essential, particularly given the global burden of albinism-associated visual impairment. Second, UKB is a population-based cohort, recruited individuals represent only milder foveal disease, and our findings may not generalise to more severe clinical presentations. Third, the zebrafish HAZ is not a true fovea, and validation in primate models will strengthen translational relevance. Finally, our multi-omics analyses are correlative and conditional studies in organoids or animal models are needed to confirm the functional roles of specific cell populations.

In conclusion, we provide a comprehensive genetic and functional framework for understanding foveal development and disease. By integrating human genomics with zebrafish functional screens and developmental multi-omics, we identify the genes and pathways governing foveal morphogenesis, demonstrate their functional relevance across species, and identify Müller glia as key early mediators of foveal specification. This work establishes a foundation for mechanistic studies and therapeutic development aimed at preventing vision loss in FH.

## Data Availability

All daIndividual-level data analysed in this study are held within UK Biobank and are available to bona fide researchers through application to UK Biobank (https://www.ukbiobank.ac.uk). All other data produced in the present study are available upon reasonable request to the authors.ta produced in the present study are available upon reasonable request to the authors

## Acknowledgements

The authors thank all volunteers participating in UK Biobank and who have made this project possible. We thank the efforts of the UK Biobank Eye and Vision Consortium for developing the ophthalmic resources that underpin this study. The authors acknowledge the help and support of staff at the University of Leicester, including the ALICE High Performance Computing Facility, the Hercules Facility, the College of Life Sciences Electron Microscopy Facility, the Division of Biomedical Services, and the Pre-Clinical Research Facility.

Supported by Wellcome Trust PhD Programme in Genomic Epidemiology and Public Health Genomics (218505_Z_19_Z); Ulverscroft Foundation Award (24/01); NIHR Senior Investigator Award to MDT (NIHR201371); Medical Research Council (MC_PC_17171); NIHR Leicester Biomedical Research Centre; EPSRC Grant (EP/X014614/1); National Eye Institute (EY022356, EY018571); Retinal Research Foundation; Research to Prevent Blindness (unrestricted grant to the Gavin Herbert Eye Institute, University of California, Irvine); and NIH Core Grant P30 EY034070 to the Gavin Herbert Eye Institute; National Organization for Albinism and Hypopigmentation; and NIAMS (R01AR077664). BPB is supported by the intramural program of the National Eye Institute, National Institutes of Health. The contributions of the NIH author(s) were made as part of their official duties as NIH federal employees, are in compliance with agency policy requirements, and are considered Works of the United States Government. However, the findings and conclusions presented in this article are those of the author(s) and do not necessarily reflect the views of the NIH or the US Department of Health and Human Services.

This work was partially supported by the National Institute for Health Research (NIHR) Leicester Biomedical Research Centre; the views expressed are those of the author(s) and not necessarily those of the National Health Service, the NIHR, or the Department of Health and Social Care. The sponsor or funding organisation had no role in the design or conduct of this research.

---

**Supplementary Figure 1:**
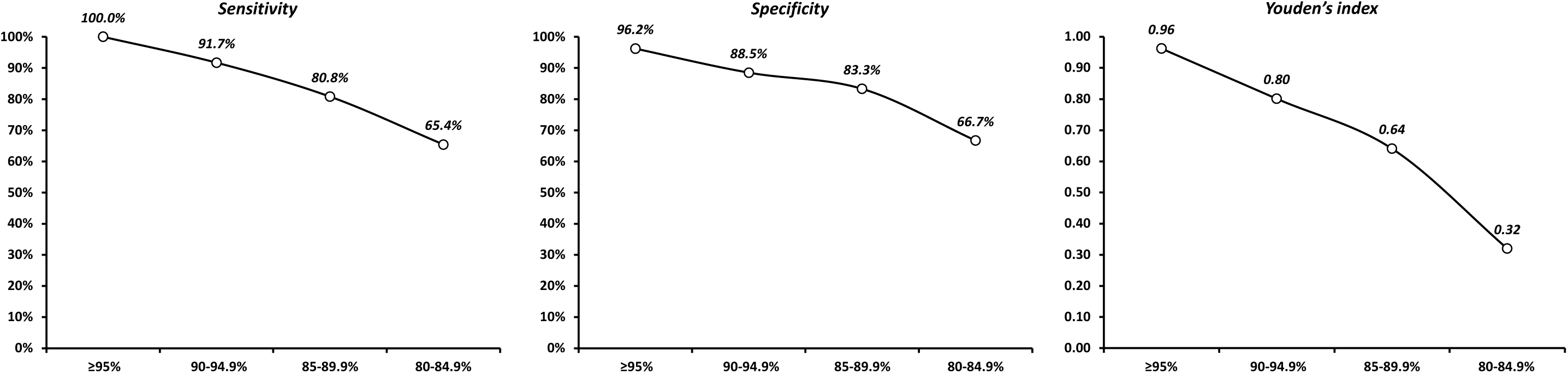
Diagnostic performance by confidence threshold. Line graphs illustrating the Sensitivity, Specificity, and Youden’s index of the model’s classifications across four confidence threshold intervals (≥ 95%, 90-94.9%, 85-89.9%, and 80-84.9%). The ≥ 95% threshold was selected for final analysis as it yielded the optimal diagnostic performance (Sensitivity: 100.0%, Specificity: 96.2%, Youden’s index: 0.96).

